# Bias in respiratory diagnoses by Large Language Models (LLMs) in Low-Middle Income Countries (LMICs)

**DOI:** 10.64898/2026.03.02.26347405

**Authors:** A. Mouelhi, K. Patel, S. Kussad, S. Ojha, A. Prayle, LMIC Medical AI Alignment Group

## Abstract

**Introduction:** Clinicians and patients are likely to increasingly use Large Language Models (LLMs) for diagnostic support. Use of LLMs mostly created in North America and Europe, could lead to a High-Income Country bias if used in Low- and Middle-Income Country (LMIC) healthcare settings. We aimed to explore if diagnostic suggestions made by LLMs are relevant in LMIC settings.

**Methods:** Five short respiratory clinical vignettes were produced. For each vignette, a group of doctors from one of 5 countries (Ghana, India, Jordan and Brazil and the UK) independently gave the 4 most likely diagnoses. 4 LLMs (ChatGPT, Claude Sonnet, Google Gemini and Microsoft Copilot) were prompted with the same vignettes. The top 4 diagnoses for each case was requested. A Virtual Private Network (VPN) was used to access the LLM from each of the 4 countries, and in a second experiment the LLM was given the same vignettes but also informed of the country in which the case was based in the prompt. The diagnoses presented by the LLMs was compared with the doctors’ diagnoses for the LMICs and also compared to the UK.

**Results:** 106 unique diagnoses were offered by 21 doctors, and 53 by LLMs with a VPN. The LLMs proposed fewer of the doctors’ diagnoses in LMICs versus in the UK - 50% (95% CI 32.6 to 67.4%) in the UK compared to 32.0% (95% CI 23.1 to 42.3%) in LMICs. This effect persisted when the LLM was informed of the location of the doctor in the prompt. Overall, LLMs performed worse in the LMIC setting (Chi-squared p = 0.028).

**Conclusion:** Doctors working in LMICs consider a wider range of diagnoses than LLMs, even when LLMs are queried from that country, or informed that they are in that country. LLMs appear to show a bias when considering likely diagnosis likely related to the epidemiology of high income countries.

## Introduction

Generative Artificial Intelligence (genAI) is emerging as a disruptive technology, impacting on almost all areas of industry. Large Language Models (LLMs) are genAI models, trained on massive corpuses of human language, which produce responses almost indistinguishable from human speech.^1^

Many LLMs are based on a Generative Pre-Trained Transformer (GPT).^2^ This LLM architecture includes a “Self Attention” mechanism which confers the ability to respond to the context within a given input prompt when outputting the next token (word or part word). This allows something similar to short term memory, and can dramatically improve outputs. LLMs include ChatGPT (from OpenAI), Claude (Anthropic) and Gemini (Google). Today, these platforms can generate responses faster than ever before, display ‘few shot’ and ‘zero-shot’ properties, and perform various tasks, such as text summarisation, creative writing, translation, reasoning and conversation.^3^ Beyond their remarkable versatility, LLMs are provided as a service by technology companies, such that many AI apps use so-called foundational models offered by large technology companies as a paid-for service. This implies that healthcare apps are subject to the same systemic biases or inaccuracies present in the host model.

The potential of LLMs in healthcare has been emphasised by their strong ability to do medical question answering, as demonstrated by GPT-4, which scored 20 marks above the pass threshold in the United States Medical Licensing Exam,^4^ an improvement over previous models.^5^ Other models have shown abilities to extract clinical data^6^ aid in medical education,^7^ automate discharge summaries,^8^ triaging ^9^ and diagnostics. ^10^ However, before LLMs can be reliably used in healthcare, several limitations need to be addressed.

When a model is trained on data from a particular region, its output may reflect that demographic group more than others. As a result, models may lack the capacity to adapt responses to different regional factors. We therefore had concerns that LLMs rely on a dataset that fails to capture global differences in healthcare. Our concerns are not unique, and we note that LLMs have been demonstrated to produce vignettes that reinforce stereotypes.^11^

The rapid progress of AI compels us to consider not *if*, but *when* LLMs can be reliably used as diagnostic aids. Given the high stakes of medical care, a major step in integrating LLMs into medicine includes identifying weaknesses to allow appropriate improvements. As bias in LLMs is a growing concern, we aimed to explore whether these models exhibit a high income country bias in medical diagnosis, particularly in respiratory medicine. By comparing the responses of models to clinical vignettes where the location of the clinical encounter could change the diagnosis, we explore this bias.

## Methods

We designed five clinical vignettes of presentations that are likely to be seen in an emergency setting. They were brief and intentionally somewhat ambiguous, such that a range of diagnoses could be considered. Additionally, we designed them such that the local epidemiology had an impact on the likely diagnosis. See Box 1 for an example vignette.

For each vignette, 4 practicing doctors from one of five countries (UK, Ghana, India, Jordan and Brazil) independently gave the four most likely diagnoses. They were identified from our own network. An online form was sent out independently to ensure no colluding. Doctors were instructed not to use any LLMs to answer. Overall, 21 doctors participated (5 from Jordan, 4 from each of the others). To ensure sufficient experience in respiratory medicine, doctors eligible for this study must have completed a minimum of two years in at least one of the following specialities: respiratory medicine, emergency medicine or general practice.

Four LLMs were prompted with the same vignettes. They were ChatGPT 4-o, Claude 3.5 Sonnet, Google Gemini-2.0-flash-001 and Microsoft Copilot GPT-4. These were the most advanced “Foundational” models available from each company at the time of the data collection in February 2025. The top four diagnoses for each case were requested.

Researchers in the UK (AM) accessed the LLMs via a Virtual Private Network which ensured that the access was from the relevant LMIC. In the first experiment, the LLMs were presented with the clinical vignettes and in the second they were also informed of the country in which the case was being seen.

The top four diagnoses from the doctors and the LLMs were then coded using ICD-11 by two researchers (AM, KP). Any disputes or uncertainties were resolved by a third researcher (AP) Initially data were aggregated into LMICs vs UK, and the diagnoses offered by the LLMs were compared to those offered by the doctors. We then explored the proportion of diagnoses offered by doctors which were also considered by the LLMs (the “coverage”) and the proportion of diagnoses offered by AI which were also considered by the doctors (the “agreement”). We undertook these analyses on both a global level, and a case-vignette by case-vignette level.

Data were using Rstudio (2024.04.1 Build748) and R (4.4.0). Data and code required to reproduce all the results and figures from this paper are available from <insert URL>. Early results from this work were presented at the European Respiratory Society Conference (Sept 2025).

## Results

5 cases were evaluated by 4 LLMs and 4 doctors in one of each of the countries Brazil, Ghana, India and the United Kingdom and 5 from Jordan. All diagnoses were coded using ICD11, which led to a total of 106 unique diagnoses offered by the 21 doctors, and 53 by LLMs with a VPN, and 59 by LLMs without a VPN but where the prompt specified the country.

The overall distribution of diagnoses in the UK and LMIC cohorts showing the overlap between LLM and doctor diagnoses is shown in Figures 1 and 2. There was a difference in the proportions of the overlaps comparing the UK to LMIC countries (Chi-squared Test: p = 0.028). The LLMs proposed 50% (95% CI 32.6 to 67.4%) of the doctors diagnoses for the UK dataset and 32.0% (95% CI 23.1 to 42.3%) of the LMICs. The wider range of diagnoses offered by doctors compared to LLMs persisted when the LLM was instructed that it was working in a specific geographical reason in the prompt (Fig 1C).

**Figure 1.**
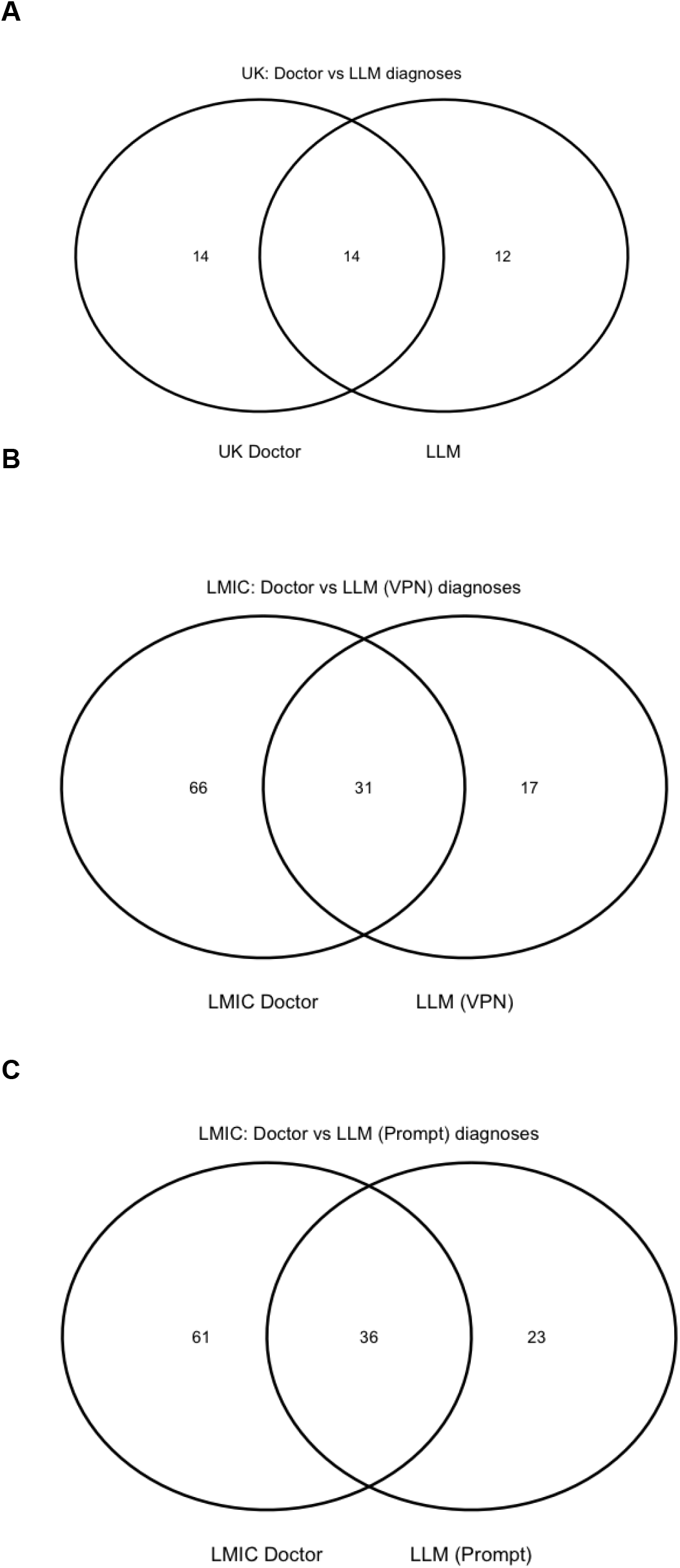
(A,B and C). The total of unique diagnoses after coding into ICD11 diagnoses were grouped by UK doctor, LLM (using a VPN for each country) for the UK (A) and for the LMICs (B) and the intersection between diagnoses is presented. Note that the LMICs present a wider range of diagnoses.

**Figure 2.**
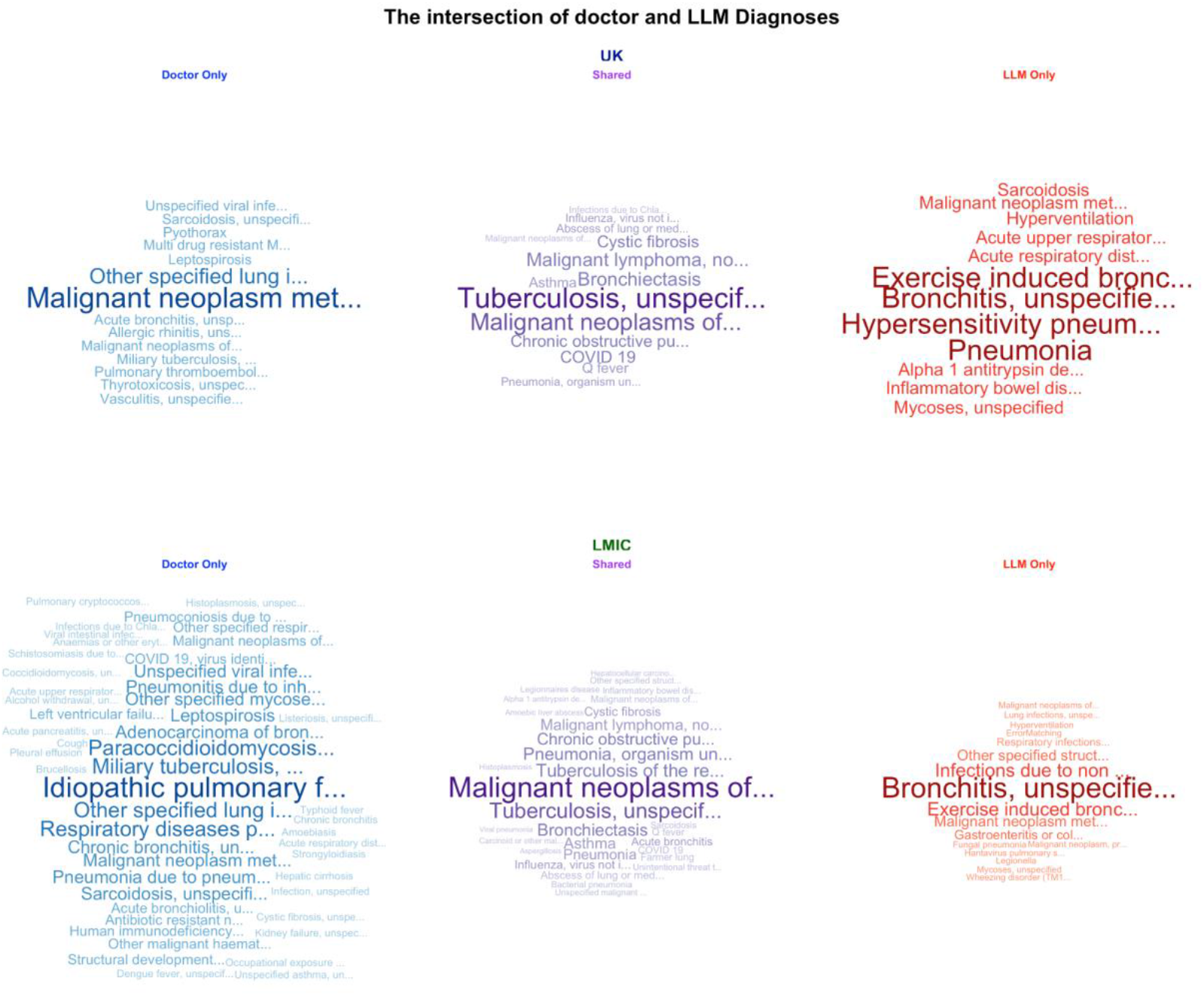
Legend: Wordclouds showing the unique diagnoses over the study. The total diagnoses were grouped by whether they were in the UK dataset (top row) vs LMIC dataset (lower row). The middle column oxford clouds shows the diagnoses considered by both the LLMs and the doctors, where as the left and right columns are doctors and LLMs only. The size of the text indicates the number of times a diagnosis was considered in a vignette. As can be seen, in the LMIC group, there were a wider range of diagnoses considered by the doctors compared to the LLMs.

We undertook further analysis comparing the agreement (the proportion of LLM suggested diagnosis that were also considered by the doctors) between LLM and doctors over the 5 cases and 5 countries (Fig. 3a), and also considering the coverage (the proportion of diagnoses considered by doctors which were also considered by the LLMs) (Fig3b). Overall, no clear patterns emerged.

**Figure 3.**
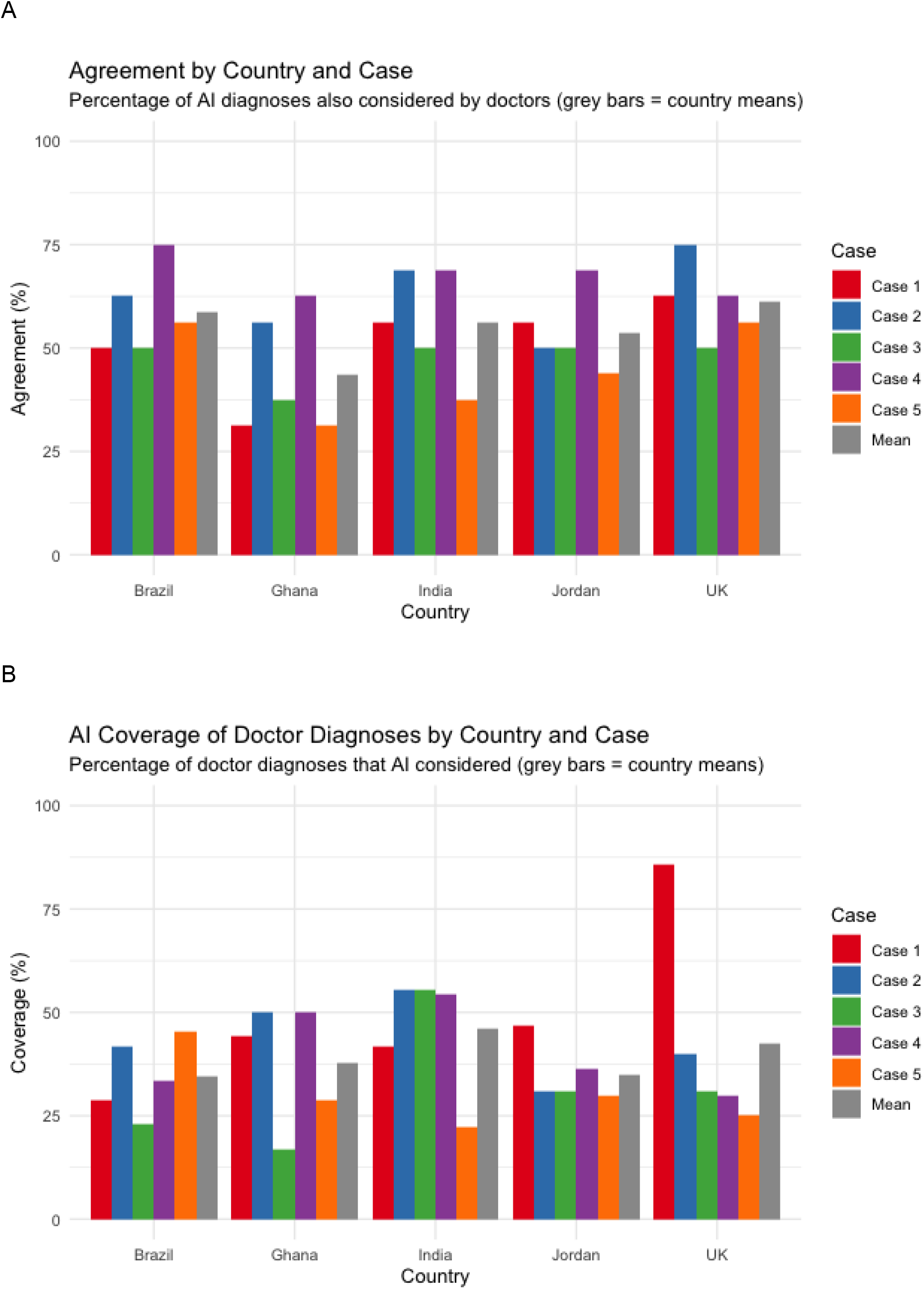

## Discussion

We have compared differential diagnoses produced by LLMs in response to a clinical vignette to practicing clinicians in an international study. Our results indicate a bias in the diagnoses offered by LLMs towards a narrower range of diagnoses (more common in high income countries). Doctors in LMICs offered a wider range of diagnoses than LLMs, likely reflecting a broader local epidemiology. Even when informing the LLM in the prompt that the clinical encounter was taking place within a specific LMIC, this issue persists. More broadly, we believe that at our work suggests that current LLMs are less aligned with the healthcare needs of LMICs compared to high income countries.

Our findings are,perhaps, unsurprising when considering how LLMs are created. The corpuses of text which form their training data likely have a high-income country bias. As LLMs and other AI technologies are likely to be used in a wide range of products in the future to aid in medical practice, this bias needs to be borne in mind. In addition to diagnosis, a role for AI has been proposed in clinical text summarisation, diagnostic radiology, histological diagnosis, and clinical noting.

It is possible that there will be similar biases in all of these, and therefore all future healthcare products using LLMs and AI should be evaluated in the setting in which they are to be used. We speculate that, for example, a clinical noting application could subtly emphasize discussion of diagnoses more prevalent in high income countries rather than LMIC when writing a clinical note, skewing the patient record.

To our knowledge, this is the first research paper investigating diagnostic bias of LLMs in LMICs. Others have investigated general diagnostic accuracy of LLMs, and at present most agree with our position that they should be used with caution, if at all. ChatGPT has been tested against 36 published vignettes from the *Merck Sharpe & Dohme (MSD) Clinical Manual*. Its average performance across all the categories was 71.8%, and whilst it achieved 76.9% on differential questions, it achieved a much lower 60.3% for differential diagnoses.^12^ This study tested more vignettes than our study but did not test the LLM against doctors’ clinical expertise and did not investigate bias. Additionally, it is possible that the vignettes presented in this manual are within the training dataset of the LLMs leading to data contamination and artificially high LLM performance.

In a randomised trial evaluating physician diagnostic reasoning, the addition of a LLM did not substantially improve clinical reasoning, with a median performance of 76% with the LLM versus 74% without.^13^ Of note, in this study, the LLM was also assessed alone, and scored 92%. This study however was limited to one high income country.

The impact of language upon differential diagnosis has been tested in a large study of over 4000 vignettes produced which represent rare diseases.^14^ This study found the performance was consistent over all ten languages, with the correct diagnosis being the top diagnosis provided by the LLM in 15 to 20% of vignettes. Notably, the languages explored in this work predominantly originate in HICs, which would not take bias in LMICs into account.

A scoping review of LLM-based disease diagnosis has suggested that inclusion of LLMs in the diagnostic pathway in partnership with clinicians’ knowledge could increase reliability, but also recognized LLM limitations such as hallucinations (i.e. essentially plausible sounding fabrications) and difficulties in handling complex diagnostic reasoning.^15^ These authors note that information gathering when producing a differential diagnosis in clinical practice is an iterative process. We agree and note that our study has similar limitations, as responses to a vignette does not adequately model clinical practice, and the complexities of clinical care are not fully captured by benchmarks such as the present work.

We reasoned that the location of the user may not be available to the LLM in the context of the prompt. Therefore, in addition to a VPN (such that the LLM website was accessed from the LMIC being tested), we prompted the LLM with the LMIC country name. All our cases are hypothetical amalgamations from our clinical experience and are intentionally vague such that the most likely diagnosis is expected to vary from one geographic setting compared to another.

We conducted this study without the knowledge, permission or communication with the technology companies responsible for the models, ensuring no changes could be made to the models which would impact performance. We note that his is also the way in which an end-user today may use the LLMs. We used the most recently developed large model at the time, assuming that this would give the best responses. We used a similar number of human clinicians to evaluate each vignette, with at least 4 repeats for each country. We used the UK as a high-income control country in order to compare the responses in a high versus low - middle income country setting. We did not evaluate or rank the individual performance of the LLMs or technology companies - our aim is not to prefer one LLM over another, but rather to make a general point about LLMs.

Our work has limitations. We did not use paid-for tiers of LLMs available at the time of the study. Whilst these may have given better responses, a large proportion of users (especially in LMIC settings) would use the free versions. Whilst we evaluated 5 countries with 5 vignettes, our sample size was small does not cover the complete range of LMICs. Our countries of choice were those where our networks allowed us to reach out to practicing doctors.

We did not explore the impact of adjusting the prompt given to the LLM to see if we could improve diagnostic accuracy (so-called “prompt engineering”), other than to give the name of the specific country. Since our initial work presented herein, LLMs have improved substantially. This is, however, the nature of AI technology, which rapidly progresses. We have not evaluated new “thinking” modes and additionally, so called “agentic” AI could help to improve the performance of LLMs with little human intervention. Alternatively, as model training data increases, it is possible that the bias may increase as more HIC information enters the model training datasets.

Therefore, despite these considerations, HIC bias may still be prevalent in newer LLMs, an area which we are currently exploring. We propose a future live benchmark, which continuously evaluates LLMs as they are released.

### Conclusions

We believe that at present, medical technologies based on AI should not be used in clinical practice in LMICs unless they have been subject to clinical evaluation in these regions. The use of genAI in healthcare is already controversial and should be more so in LMICs due to the bias which we have found.

We suggest that companies and other organisations developing healthcare related genAI products or services rigorously evaluate their offerings in all geographical regions which they are available, prior to use in routine clinical practice. The developers of foundational models should assess their models for LMIC bias at the time of their model release.

Finally, we propose the development of a new benchmark study for training models to assess healthcare based models for bias against LMICs.

## Box 1. One of the 5 cases

A 45-year-old male comes into the emergency department with weight loss, a fever of 38.5C and dehydration. They are very concerned as they report that they have had mild, intermittent episodes of blood-tinged diarrhoea. These symptoms have been persistent for 3 months. He has a history of anorexia. Examination revealed an enlarged liver and mild tenderness when palpated. There were some crackles heard in the left lung. A CT scan was carried out and it revealed pleural effusion as well as a mass in the left lower lung lobe. They have a history of smoking 15 a day for 20 years and drink 5 units of alcohol a week. What are the top four most likely diagnoses?

## Data Availability

Data are available on request. We are currently depositing in an on-line repository and will update this information when this is available.

## Appendix 1 LMIC Medical AI Alignment Group Authors

The following are members of the Medical AI Alignment Group and are considered co-authors.

Alba Pereira

Suellen Soares de Sousa

Flávyus Luciano Cardoso

Ricardo Amorim Correa

Samuel Twumasi

Samuel Amo-Mensah

Boston Frimpong

Padmond Badu Agyemang

Dia’ Aldeen Suhel Mohammad Suleiman

Farah Taisir Khalil Alsheikh

Mahmoud Hussein Ahmad Mohammad

Dana Ahmad Suleiman Elmughrabi

Shalini Ojha

Aditi Agarwalagarwal

Lubna Makhija

Preetha Padmanabhan

Joanne Ollerton.

John Merry

Owain Powell

Alistair Ramsay

## Notes

### Competing Interest Statement

No authors or their institutions have received their institutions received any payments or services from a third party in the past 36 months with entities that could be perceived to influence, or that give the appearance of potentially influencing, the submitted work.
Andrew Prayle declares grant funding from Vertex Ltd paid to his institution and support to attend an international conferernce.

### Funding Statement

This study did not receive any funding.

